# Experiences of delivering social homecare at end-of life: insights from a qualitative study drawing on multiple perspectives

**DOI:** 10.1101/2025.07.11.25331358

**Authors:** Caroline White, Cat Forward, Zana Bayley, Helene Elliott-Button, Justine Krygier, Alison Bravington, Samina Begum, Joan Bothma, Jamilla Hussain, Miriam Johnson, Colin Moss, Mark Pearson, Helen Roberts, Paul Taylor, Jane Wray, Liz Walker

**Affiliations:** Wolfson Palliative Care Research Centre, Hull York Medical School, University of Hull; Health and Social Care Workforce Research Unit, Virginia Woolf Building, Kings College London, WC2B 6LE; Faculty of Health Sciences, University of Hull; Yorkshire Quality and Safety Research Group (YQSR), Bradford Institute for Health Research, Bradford Teaching Hospitals NHS Foundation Trust, Bradford; Involve Hull, Institute for Clinical and Applied Health Research, University of Hull; Cera Care, Essex; Bradford Teaching Hospitals NHS Foundation Trust; Institute for Clinical and Applied Health Research, University of Hull; Sheffield Centre for Health-Related Research, University of Sheffield, and St Luke’s Hospice, Sheffield, SN11 9NE

**Keywords:** Social care, homecare, end-of-life care, loss and bereavement

## Abstract

Social homecare workers (HCWs) play an important role in supporting people with care and support needs who wish to remain at home as they approach the end of life. However, the experiences of these HCWs have been neglected within policy and research, leaving gaps in knowledge regarding the challenges they face, and the support needed. Given the difficulties in recruiting and retaining staff in the home care sector, a better understanding of the experiences and needs of this workforce is essential.

This paper reports on the findings of a multiple case study, using semi-structured interviews carried out with HCWs, managers, clients, carers (families/friends), health and social care practitioners and service commissioners. Interviews were supported by Pictor, a visual elicitation method used to map networks and relationships. 133 individuals participated across three sites in England chosen to reflect different demographic characteristics. Data were analysed using reflexive thematic analysis in NVivo 14.The findings presented here highlight three themes in respect of HCW experiences: the unique privileges and challenges of providing care at end-of-life; the relational aspects of care important at end-of-life; and the multi-agency challenges and opportunities experienced at end-of-life, with the wider factors such as policy and the care sector environment which can influence HCW experiences at work also considered.The findings are discussed in the context of an adaptation of Bronfenbrenner’s Ecological Systems Theory to explore the different levels operating in the community care network, and are related to current evidence with suggestions made for policy, practice and future research.

## Introduction and background

Many people’s preference is to die in their own home, with a trend towards increasing numbers of deaths at home in many nations (Higginson et al., 2013; Bone et al., 2018; Funk et al., 2023). However, a home death is a ‘distant reality’ for many, because of social inequalities and lack of access to appropriate support which impacts on location and experience of death (Higginson et al, 2013, p.918; Funk et al., 2023).To facilitate the choice to die at home a support network including family and friends (hereafter referred to as carers) and health and social care practitioners is required (Gomes and Higginson, 2006; Bone et al, 2018). Social homecare workers (HCWs) are an essential element of this network, often providing practical assistance such as personal care and medication administration, alongside emotional support, to the person dying and their carers, although commissioning is usually restricted to practical support (Forward et al., 2024). Carers may experience end-of-life care as emotionally and physically demanding and financially costly; the finding that 9% of carers providing personal care to a relative or friend at end-of-life would not do so again under similar circumstances highlights the need for better support both for those dying and their carers (Ewing et al., 2012; Johnson et al, 2016; Mogan et al, 2024)

In England, where this study was located, most homecare is positioned within the social care sector, alongside but separate from healthcare, and operating within a ‘mixed economy’, in which care is primarily delivered by independent providers including private companies and charities, with a smaller amount delivered by agencies within the statutory sector (local authorities and health) (Homecare Association, 2024). Care is commissioned by local authorities, who adhere to stringent eligibility criteria, and by those who self-fund or ‘top up’ their care costs (Homecare Association 2024). A recent report has indicated around 420,000 people are employed in the homecare sector providing direct care, representing a sizeable workforce (Skills for Care, 2024).The English social care sector has long been viewed as in crisis, due to sustained under-funding as a result of UK government policies and an imbalance between the care supply and demand (Glasby et al., 2021; Burns et al., 2022); similar challenges have been reported elsewhere, reflective of strained long-term care sectors internationally (Deusdad et al., 2016; Bennett et al., 2020; Gori, 2019; Scales, 2021; Zagrodney et al., 2023; Bergvist et al, 2024)

The stretched resources within homecare result in challenging working conditions for HCWs who are frequently poorly paid and are often required to meet costs of travel to clients, as well as to provide care in their own time if calls overrun (Bottery et al., 2018;Timonen and Lolich 2019; Ravalier et al., 2019; Burns et al., 2023).Their working hours may be unpredictable: 42% are on zero hours contracts, wherein they lack the assurance of minimum hours of employment (which may be the preferred option for some, but often creates precarity), while others may be required to work extended hours if agencies are short-staffed (Ravalier et al., 2019; Skills for Care, 2024; Burns et al., 2024). Further, many care calls are of short duration, often separated by long gaps for which workers are unpaid (Ravalier et al, 2019). HCWs appear to be held in low esteem (Bottery et al., 2018, Bergqvist et al., 2024); homecare agencies struggle to recruit and retain staff, often relying on international recruitment, with high levels of staff turnover at around 25% (Skills for Care, 2024).The wider homecare market is volatile, with high turnover of agencies, and significant levels of business failure (Bottery et al., 2018; Skills for Care, 2024).These factors risk low staff morale, loss of knowledge and experience from the sector due to staff attrition, and impact on agencies’ abilities to provide the continuity of care required by clients and carers at end-of-life (Devlin and McIlfatrick, 2010; Manthorpe et al, 2019).

There has been little research on the experiences of HCWs providing care at end-of-life, and the support and training which could facilitate this role (d’Astous et al, 2019, Forward et al., 2024).This mirrors the scant policy attention to their contribution in facilitating care of the dying at home (Author’s Own (a) – in preparation). Existing literature (Forward et al., 2024) highlights the complexities and challenges of providing homecare at end-of-life.These include recognition of the emotionally charged environment in which HCWs support others’ emotions; their isolation within the wider care network; and their lack of preparation and training for their role in providing end-of-life care.

The SUPPORTED study aimed to inform improvements in the quality and sustainability of homecare at end-of-life, through exploring the experiences of HCWs and developing support and resources to address training gaps (Bayley et al., 2023).

## Methodology

The study protocol is detailed elsewhere (Bayley et al, 2023) but summarised here.We used a pragmatic paradigm: the 3-stage DESCARTE (Design of Case Study Research in Health Care) case study model (Carolan et al., 2016) employing a multiple case study approach across three sites in England.This allowed diversity in regional resources and practices to be recognised and enabled comparison between areas, supporting robust applicability (Paparini et al., 2020). A qualitative approach was used to best investigate the previously under-explored experiences of HCWs at end-of-life, informed by multiple perspectives and subjectivities.

### Recruitment

Study sites were selected to reflect variation in local population demographics, ethnicity, and socio-economic profiles.These included Site 1, a Northern city with a diverse ethnic population and significant levels of poverty, although with pockets of affluence in the surrounding, more rural area; Site 2, a Southern borough, which was relatively affluent, with an ageing population, and relatively low ethnic diversity; Site 3, a Northern city with high indices of multiple deprivation and in which the substantial majority of the population was identified as White British. Participants were recruited using a form of purposive sampling to enable inclusion of diverse experiences, with, for example, emphasis placed on including HCWs and managers from agencies which advertised the provision of end-of-life care, and those which advertised generic homecare only. Recruitment was conducted through homecare agencies and hospices, social media, existing networks, and with support from the relevant Regional Research Delivery Networks, and through snowballing, in which participants forwarded information to others in their networks. Inclusion and exclusion criteria were developed for each participant group (Bayley et al., 2023) and are summarised in the Appendix. Participants were provided with Participant Information Sheets and could ask questions prior to giving written consent to participate. HCWs, clients and carers were offered shopping vouchers in recognition of their contribution to the study.

### Data generation

In-depth interviews were conducted at a time and place convenient to participants (in-person or online). Participants were invited to undertake a Pictor-guided interview (King et al., 2013), a graphic-elicitation method which utilises arrow-shaped sticky notes enabling participants to map out those involved in an episode of care or particular event, illustrating care networks and relationships.This method has been used successfully in studies exploring care provision in palliative care and other settings to elicit in-depth insights into potentially complex care scenarios and relationships (Bulk et al., 2020; Bravington et al., 2024). Participants could undertake a semi-structured interview if preferred.

Interview guides were developed for each participant group by the research team with support and guidance from the two study advisory groups (service users; HCWs).These were used to guide interviews whether or not participants elected to construct a Pictor chart. The main questions for each participant group are provided in the Appendix. Interviews were conducted between May 2023 - May 2024, by [withheld during review]. All were audio-recorded, transcribed and anonymised prior to analysis.

### Data analysis

Reflexive thematic analysis was undertaken (Braun and & Clarke, 2022) including familiarisation, coding, development, refinement and definition of themes.The data analysis was conducted by [researchers anonymised during review] with regular input from the wider research team who collectively had backgrounds in social work, social care (including homecare), palliative care, occupational therapy, psychology, nursing, and lived experience as carers.The researchers each read a sample of the transcripts to immerse themselves in the data and met with members of the wider team to bring insights to code development, taking an inductive approach. Coding of the data was undertaken using NVivo 14, following the process described by Braun and Clarke (2019; 594) which involves *a continual bending back on oneself – questioning and querying the assumptions we are making in interpreting and coding the data,* as different understandings and interpretations of the codes emerged among the team, requiring refinement of codes. Themes were generated following an iterative process in which related codes were clustered into working themes, which were refined and revised, then named to capture key meanings.The final themes and constituent codes were also reviewed by our study advisory groups and two public members of the research team, to ensure whether researcher interpretations resonated with their experience.

The analysis also drew upon Bronfenbrenner’s Adapted Ecological Systems Theory (Pask et al., 2018) to explore the complexity of homecare at end-of-life, and the systems which inform HCW experiences.This model considers the HCW, their needs and circumstances (microsystem); their interactions with clients, carers and practitioners (mesosystem); the wider context of multi-disciplinary end-of-life care services within the locality (exosystem); the social/political context (macrosystem); while also highlighting the dynamic influence of time in end-of-life care at all levels (the chronosystem).

Pictor data were analysed following King et al. (2013), utilising aspects of the Pictor charts such as direction and spacing of arrows, and frequency counts of professional disciplines or role patterns across the charts.

Additionally a matrix analysis was undertaken to compare the similarities and differences at the three case study sites.The data for each theme and sub-theme was then analysed at site level; data within each theme was read and a summary created for each site, and exemplar quotes provided, enabling the identification of any key site level differences to be identified.

### Ethics

The study received ethics approvals from the West Midland (Coventry and Warwickshire) Research Ethics Committee (reference 32/WM/0030, 31^st^ March 2023). Key ethical considerations related to the emotional impact of participation, with client and carer criteria excluding those expected to die within the following seven days or who had experienced bereavement during the previous three months. Illustrative participant quotes are included, with any identifying detail redacted and codes given following the format: site number-participant group-participant number (e.g. 1-HCW-1).

## Findings

### Participants

133 interviews were conducted. Participants included HCWs (41), managers (22), clients (18), carers (20), health and social care practitioners (29) and commissioners (3).

Participant characteristics are shown in Table 1.

**Table 1.** Demographic characteristics of participants.

|  | HCW | Manager | Client | Carer | Commissioner | Practitioner | Totals |
| --- | --- | --- | --- | --- | --- | --- | --- |
| **Site** |  |  |  |  |  |  |  |
| 1 | 12 | 8 | 4 | 3 | - | 7 | 34 |
| 2 | 12 | 6 | 8 | 10 | 2 | 11 | 49 |
| 3 | 17 | 8 | 6 | 7 | 1 | 11 | 50 |
| **Totals** | **41** | **22** | **18** | **19** | **3** | **29** | **133** |
| **Ethnicity** |  |  |  |  |  |  |  |
| Asian/Asian | 3 | 1 | - | 1 | - | 1 | 6 |
| British | 1 | 3 | - | - | 1 | - | 5 |
| Black/Black | 1 | - | - | - | - | 3 | 4 |
| British | 4 | 3 | - | - | - | 2 | 9 |
| Mixed | 1 | - | - | - | - | 2 | 3 |
| Other | 28 | 15 | 18 | 18 | 2 | 20 | 101 |
| White Other | 38 | 22 | 18 | 19 | 3 | 28 | 128 |

| White British |  |  |  |  |  |  |  |
| --- | --- | --- | --- | --- | --- | --- | --- |
| Totals |  |  |  |  |  |  |  |
| Age |  |  |  |  |  |  |  |
| 18-20 | 1 | - | - | - | - | - | 1 |
| 21-30 | 8 | 3 | - | - | - | 1 | 12 |
| 31-40 | 11 | 2 | - | - | - | 8 | 21 |
| 41-50 | 8 | 6 | - | 1 | 2 | 10 | 27 |
| 51-60 | 8 | 7 | 1 | 8 | - | 7 | 31 |
| 61-70 | 2 | 4 | 5 | 5 | 1 | 2 | 19 |
| 71-80 | - | - | 5 | 3 | - | - | 8 |
| 81-90 | - | - | 5 | 2 | - | - | 7 |
| 91+ | - | - | 2 | - | - | - | 2 |
| Totals | 38 | 22 | 18 | 19 | 3 | 28 | 128 |

\*5 participants did not disclose ethnicity, 5 did not disclose age, affecting totals.

Eighty-nine interviews took place in-person and 44 online. Forty-three participants (32%) undertook a Pictor guided interview; reasons participants declined to construct a Pictor chart included interview conducted online; participant visual impairment; personal preference (e.g. participants perceived themselves as ‘not a visual person’). Interview duration ranged from 7 – 100 minutes (average 37 minutes).

Multiple homecare providers were contacted across each site. Most participating agencies were independent. Some offered generic homecare, others advertised specialist end-of-life care provision. One agency (Site Three) was based within a provider of statutory healthcare and provided short term end-of-life care, often in the final days and hours of life.

Through the reflexive thematic analysis, five themes were generated with sub-themes.The first three themes are discussed in this paper and are summarised in Table 2. Themes in respect of training and support needs, and inter-professional collaboration are reported elsewhere (Author’s Own – in-preparation b); Authors’ Own, under review).

**Table 2.**
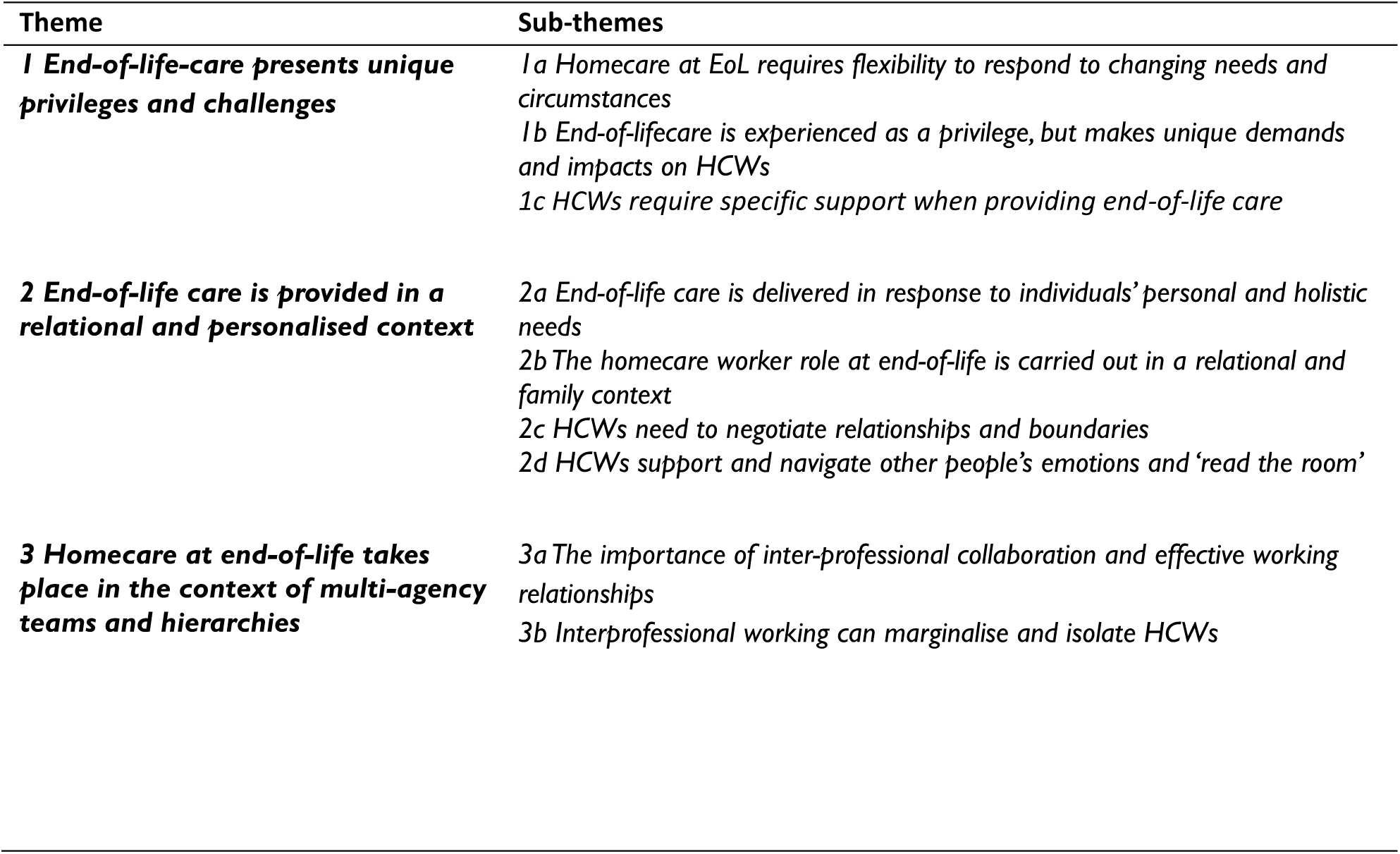
Themes and sub-themes.

### Theme 1 – End-of-life-care presents unique privileges and challenges

#### Homecare at end-of-life requires flexibility to respond to changing needs and circumstances

As clients approach end-of-life, their deterioration can be rapid and unpredictable, requiring care to be responsive and flexible to meet individuals’ changing needs. HCWs were required to undertake new and specific tasks such as managing pain medication and new equipment, and to adapt care to accommodate changing mobility, pain, and dietary needs, and changing personal preferences and wishes. This required thoughtful tailoring of care plans to respond to in-the-moment changes, and ensure evolving needs were prioritised, with ensuring client comfort often the primary concern for HCWs as clients approached end-of-life.Thus, care plans were followed flexibly, with personal care provided in-line with client preferences and at the time they could best tolerate this. HCWs and managers reported that tasks could be omitted or postponed and that providing emotional support to clients and carers was often perceived as more important than delivering personal care at a prescribed time:

> *You have to change your care sometimes hourly with a person that’s dying….So we’re…keeping ‘em clean and dry and blah-blah-blah, but if they’re literally end of life, why are you gonna pull that person around if they’re in pain? So the things that are really important in normal day-to-day care…feeding, drinking….they become unimportant…what’s important is their comfort, so…your care changes (2-HCW-2)*.

HCWs also needed to adapt their communication as individuals’ preferences and abilities changed.

This included an increased focus on being attentive to non-verbal communication and cues.

Managing visit times was important to enable HCWs to respond flexibly to clients’ changing needs, for example, allowing HCWs to stay longer in response to changes or crises, and avoiding rushed care, which was important to clients, carers, HCWs and managers alike. Agencies which did not provide strictly timed calls and could rearrange rotas to enable HCWs to stay with clients when necessary, facilitated this flexibility. However, this also meant that clients, carers and practitioners were unable to predict when HCW visits would occur, and practitioners were unable to plan their visits to coincide with those of HCWs.

#### End-of-life care is experienced as a privilege but makes unique demands and impacts on HCWs

HCWs emphasised their commitment to providing good end-of-life care.They reported deriving satisfaction and pride in supporting clients to die with dignity, at home, in company and comfort, and without pain:

> *If you’re the last person that’s listening to Roy Orbison [a singer] with ‘em, if you’re the last person that cooks ‘em a meal, if you’re the last person that sees ‘em and makes ‘em comfortable…it’s that rewarding (3-HCW--10)*.

While experienced as a privilege, end-of-life care also confronted HCWs with uncertainties, loss of important relationships, grief and bereavement, and the need to manage endings.Together these highlight the emotional toil associated with end-of-life care, and required HCWs to manage such uncertainty. Determining when individuals are close to death is difficult and HCWs experienced shock when clients died more quickly than anticipated.They experienced uncertainty on arrival, feeling unsure whether the person would still be alive, and when leaving, recognising this could be their final visit:

> *The stepping out of my car and walking towards their door and…the unknown of whether or not they’ve gone yet…that’s the challenging part for me (1-HCW-8)*.
>
> *It can be really hard, cos every time you leave ‘em are, are you gonna come back to them? You… don’t know, your heart’s always like are they gonna be there? When the phone bleeps….is it them, is it not? (I-HCW-1)*.

HCWs experienced distress at witnessing individuals’ declining health, being present at, or ‘first on the scene’ after death. Further, when clients died, HCWs could experience distress, although the impacts appeared, in part, determined by the duration of their relationships:

> *She ended up with a terminal cancer diagnosis and where she’d already been our client for…two years; and I think when that happens, when you’ve gotten to know them and they get diagnosed like that, that’s tough, that’s really tough, as opposed to them coming directly to you as a palliative care client (1-Manager-1)*.

Care provision at end-of-life extended to provision of care for the person’s body after death:

> *We had two night staff on and the two day staff that were supposed to come on actually came and then the four of ‘em all got her changed and…..made her look…..she actually did look beautiful, (3-Manager-4)*.

HCWs appeared to experience the death of their clients as especially distressing when clients were very young, had children, or where the relationship was close and/or long-standing, when HCWs had recent personal experience of bereavement or were encountering death for the first time, for which some had not been well prepared.There was variation in respect of the emotional impact of client death, with individual differences among HCWs and variation in the impact of different deaths.

However, it was evident from participants’ responses that even when they felt they had become accustomed to death in their professional lives and developed coping strategies, some deaths were especially painful and the loss of relationships with clients and families could be hard-felt, akin to a death in the family, and the sense of loss could be enduring. Participants often recognised the need to ‘be professional’, ‘carry on’ and fulfil their responsibilities to other clients:

> *It is upsetting but you put it in a box and you close that box and you carry on, because it’s somebody else that needs yer (1-HCW-11)*.

However, this meant that emotions might be masked while at work, with potential for the impact on individuals to go unnoticed.

Managing endings was an important element of end-of-life care. Attending funerals provided an opportunity to mark the loss and honour the person who died; this was important for some HCWs, and it could be painful if families did not want them to be present. However, some preferred not to attend, and to mark the death in their own way:

> *I have been invited to funerals…I always say no, but I always make sure when…the hearse goes to the church, when they carry the coffin in, my car is parked somewhere where I can see it….that’s like my way of saying goodbye (1-HCW-1)*.

Maintaining contact with families after client death could also be important and valued both by carers and HCWs. However, this required a careful balance in respect of boundaries and what was appropriate within professional relationships. Some HCWs and managers were clear that contact after death, while important, had to be carefully managed and/or time limited; some managers however recognised the duration of relationships and acknowledged that they:

> *Turn a blind eye to the fact that the relationships…are not quite as professional after that length of time as they’re supposed to be (1-Manager-2&3 – joint interview)*

Thus, in some instances, HCWs appeared to have maintained enduring contact with families, sometimes long after clients died.

#### HCWs require specific support when providing end-of-life care

HCWs used a range of strategies, drawing on personal and work relationships to enable them to cope when clients died. Support from managers and the wider care organisation was critical and is explored elsewhere (Authors’ Own (b) in preparation).This included managers recognising that not all HCWs feel able to support end-of-life care, and that some require a break from providing such care after a bereavement within or outside work. However, rearranging rotas to accommodate this could be challenging, and while managers highlighted their commitment to this, the following comment also suggests that staff might be under pressure in some organisations to use their own resources following a bereavement:

> *We…give them a break..before asking them whether they…are ready to go to another client….they put in for leave (1-Manager-1)*.

Further, it was noted that how HCWs are informed of client death is important, and should be done with sensitivity, rather than through communication methods such as group chats.

### Theme 2 - End-of-life care is provided in a relational and personalised context

#### End-of-life care is delivered in response to individuals’ personal and holistic needs

Participants recognised that clients at end-of-life could be distressed and frightened, experience indignities as health failed, and made sometimes unwelcome transitions to accepting intimate and personal care:

> *I tell you what, it’s…awful to, having to expose your body to a young woman (sighs)(2-Client-6)*.

Therefore, HCWs sought to make the experience of receiving care as dignified and acceptable as possible.This included finding ways to support the person to be as independent as they wished, stepping in when support was needed, and working to help the person feel as physically and emotionally comfortable as possible, rooted in an understanding of their individual preferences:

> *One lady liked….Louis Armstrong…so what we used to do was…have our mobile phone playing Louis Armstrong in our pocket….because at that point she was quite uncomfortable being moved; whilst one was doing the personal care the other one was either stroking to her, singing together, she may hum or she might move her fingers and tap to the music….that made things for the personal care much easier for her, and more relaxed (2-Manager-4)*.

HCWs and managers highlighted the importance of the care encounter as a social experience as well as one in which practical care tasks were undertaken, recognising that HCWs were often among those spending most time with clients, some of whom were lonely, and the value of humour and ‘banter’ were often noted. Further, they worked to ensure that clients experienced pleasure through addressing the *little things (1-Manager-7)* that mattered to them. HCWs gave numerous examples of the ways they enabled clients to ‘live while dying’, based on their knowledge of the person:

> *We had a lady who wanted….to just get out one last time to go and sit in a coffee shop and watch the world go by.We managed to get that sorted through equipment services, getting her a wheelchair, and we got that and…. she died the next day (3-Manager-5)*.

Attending to clients’ cultural and religious beliefs was also identified as important, for example recognising the importance of same-gender care within some cultures and ensuring that after death the person’s body was handled appropriately and in-line with cultural and religious beliefs, highlighting the importance of:

> *Knowing those things in advance….knowing things like…the religious needs of a service user, because they vary depending on, not only the religion of somebody but…..how people want to adhere to that religion….So it’s about knowing that sort of information and being able to support the service user (3-Manager-3)*.

Ensuring that the HCWs allocated to clients would be able to connect with and get to know them, and whose communication style matched individuals’ perceived preferences was identified as an important role for managers:

> *I can just think of some of our carers that, with the best of intentions…would try and be a bit too jolly….you’ve got to pick your carers (3-Manager-8)*.

Moreover, participants highlighted difficulties and tensions when carers, clients and HCWs did not share a common language:

> *Neither of them had English as their first language, and nor did the patient….they didn’t share a common language; so they all (sighs) it struck me that…they were doing everything that they could but it was so limited because there was a vital bit of information that…neither party could sort of get across to the other party cos they couldn’t really speak to each other (2-Practitioner-11)*.

Examples were given of HCWs using translation apps and learning key words from family members, to bridge linguistic gaps, but the potential for lost information, as well as a negative impact on building relationships and trust was evident.

#### The homecare worker role at end-of-life is carried out in a relational and family context

The importance of relationships when supporting clients at end-of-life was highlighted, and these were valued by HCWs, clients and carers alike. Relationships were grounded in HCW knowledge of the person, which enabled them to deliver personalised care, understand how to best encourage the person or help *lift the mood (1-Practitioner-7)* and to recognise change or signs that further support might be required.

HCWs frequently also interacted with carers (although not all clients have supportive family networks), providing care together or separately.They also provided support directly to carers, responding to the needs of co-resident carers and those living elsewhere.This support was seldom on the care plan and had to be managed within the time allocated for the visit. HCWs viewed their role within a ‘whole family’ system, rather than focusing exclusively on the needs of the client:

> *You’re there for everybody and that’s what we say to ‘em “We’re not just there for the client we’re there for you as well.We’re that sorta like stepping stone if you just want a chat” (3-HCW-10)*.

HCWs identified important roles in providing emotional and practical support, advice, information and updates to carers, who were often experiencing end-of-life for the first time.Their experience often equipped them to notice when clients were approaching death, but required them to navigate dilemmas in which they did not feel qualified to answer families’ questions or share their insights, which were often outside the scope of their roles:

> *I’ve seen quite a few deaths and…I can tell…when someone’s about to die…but…I can’t tell the family that cos I’m not a doctor…I would never say to the family…“I don’t think they’ve got long left.” But you…see it, cos you notice the things that are happening (3-HCW-17)*.

Although HCWs frequently reported working to provide support to carers, some gave examples of negative experiences which are likely to impact on relationships:

> *There was only two people in that company that ever came that knew how to care for somebody that was ill and understood (3-Carer-3)*.
>
> *The end of life was raw egg, it was the washing, it was the poo everywhere, it was the dirty clothes, it was not taking tablets, taking the wrong tablets (2-Carer-4)*.

#### HCWs need to negotiate relationships and boundaries

Homecare provided at end-of-life is physically and emotionally intimate and delivered within the privacy of the domestic space. Close relationships could be forged, which were often perceived as akin to those of families and friends. HCWs often went beyond the roles expected within purely professional relationships, with some visiting outside scheduled visits, providing treats or food paid for with their own money, undertaking tasks outwith the care plan, and sharing personal phone numbers, although *as carers we’re not actually meant to give ‘em our telephone numbers – most of us do (2-HCW-3).* The development of friendly, familial relationships appeared to enable HCWs to transcend purely instrumental care and build trusting relationships which made care at a difficult time more bearable. However, what constituted appropriate relationships in this context appeared complex and sometimes contradictory:

> *We always say to them about being professional, professional boundaries….but we want ‘em to be human as well (1-Manager-4)*.

HCWs were therefore required to walk a complex line in which boundaries could be easily crossed, sometimes with difficult consequences. Clients perceiving HCWs as ‘like family’ could be upsetting for their relatives, and there were examples of both HCWs and clients overstepping boundaries or behaving in ways which were considered harmful.

While there were many examples of positive relationships, there was also potential for conflict, competing priorities, and exposure to difficult family dynamics and distress. HCWs and carers could have different perspectives about what was in clients’ best interests, carers could lack understanding of the parameters of HCWs’ roles, and carers could be concerned that treatment or treatment withdrawal may precipitate death. In these circumstances HCWs could be under pressure to carry out tasks beyond their remit:

> *I’ve had some confrontations because of that [client nil by mouth], they’re like “Well that’s not enough, they need water, they need to be able to drink” because… they don’t agree with the doctor and they don’t agree with it’s nil by mouth now, and they don’t actually understand (2-HCW-3)*.

The often-negative reputation of homecare (Bergqvist et al., 2024), coupled with family concerns at this time of heightened anxiety meant that carers sometimes installed monitoring devices or were present during client care; this could make HCWs feel uncomfortable and ‘under surveillance’, while appreciating carer concerns, and client dignity also had to be considered.

#### HCWs support and navigate other people’s emotions and ‘read the room’

Providing care at end-of-life required HCWs to be aware and supportive of client and carer emotional needs, while also supporting colleagues.They recognised that at end-of life their behaviour and demeanour might need to be momentarily adjusted in response to client and carer needs.This required them to make nuanced judgments about individuals’ needs, and the ability of HCWs to use intuition and ‘read the room’, working out from often unspoken signals what was required was highlighted:

> *You don’t know when you’re walking into that room who has been there before you. Macmillan [cancer charity funded nurse specialist] might have been, broken some obviously bad news….family might have obviously had some tensions between each other obviously with it being emotional. So you just have to kind of approach with kind of a bit of caution…. just kinda gauge the vibe from there (3-HCW-5)*.
>
> *Staff need to know and need to be aware of …pick up on the cues…that they need to actually just step out…to allow the family and the loved ones to have that time, but know when to step back in, in the event that there’s something sorta specifically required (3-Manager--3)*.

They were also aware that sometimes individuals needed ‘normality’ and *don’t want you to be in there quiet and moping about (1-HCW-1).* Therefore, flexibility and tailoring their communication and interactions to clients’ changing preferences appeared an important element of end-of-life emotional care. However, some appeared uncomfortable or fearful of discussing end-of-life with clients and sought to ‘smooth things over’:

> *Oh she tells yer “I’m not gonna be here for long”……What can you say? I don’t think anything prepares yer for that sorta conversation (1-HCW-12)*.

This highlights an important area in which HCWs could be better prepared and supported.

The delivery of personalised and relational care to clients and carers was valued by all participant groups. However, this was impacted by time and the frequent requirement to provide care in the context of short calls in which HCWs were required to make timely visits to other clients. Services which did not operate strictly timed calls or where longer visits were commissioned appeared to more easily facilitate personalised and relational care:

> *That’s what I love about this service….we’re never rushed….we get so much time to spend and it’s not like, right, you’ve got fifteen minutes, we need to get this and this done and then we’re out; we always get time to, to get to know the clients….we build up a friendship…(3-HCW-9)*.

### Theme 3 - Homecare at end-of-life takes place in the context of multi-agency teams and hierarchies

#### The importance of interprofessional collaboration and effective working relationships

Interviews and Pictor charts demonstrated the range of practitioners involved in end-of-life care; these included community and Macmillan nurses, occupational therapists, General Practitioners (family doctors), physiotherapists, speech and language therapists, community palliative care services, charities such as hospices, and dieticians. HCW experiences of inter-professional working appeared variable and unpredictable:

> *Sometimes it goes swimmingly well and sometimes it goes absolutely terribly…and there’s no guarantee which one you’re gonna get (2-HCW-2)*.

There were examples reported of supportive and collaborative practice, in which information was shared effectively and HCWs felt well supported and informed by practitioners and *part of the team (3-HCW-8)* with positive working relationships reported:

> *The district nurses are really good…they’re really reassuring and they just go through everything step by step (1-Manager-6)*.

However, HCWs and practitioners also appeared to experience barriers which hindered inter-professional working. HCW and practitioner visits often occurred at different times, but when present at the same time they could be engaged in separate activities with limited interaction, so were simultaneously together, yet separate: *we don’t work together…we’re just there at the same time doing different roles (3-HCW-1)*.

Communication systems could also be fragmented and the information available to HCWs limited. In some instances, practitioners were perceived as unwilling to discuss clients’ needs with HCWs, to share information or they were omitted from discussions. Important information was not always communicated, with the consequent risk that staff may not be equipped to meet specific needs:

> *You get the poorest paperwork you’ve ever seen with an end of life patient….sometimes it’ll tell yer what they’re dying of, sometimes it doesn’t, sometimes it don’t hardly tell you anything and you get there and they’ve got a PEG [abdominal feeding tube], they’ve got a catheter, but [the paperwork] hasn’t told you that (3-Manager-4)*.

HCWs usually lacked access to the systems used by healthcare practitioners. An exception to this was a homecare agency based within a provider of statutory healthcare in site 3, which had access to NHS systems, allowing HCWs to view patient records and easily contact healthcare practitioners, share information and request support.Where HCWs kept notes on digital systems these were inaccessible to healthcare practitioners, in contrast to previous experiences of paper records which were more widely accessible between services because they were often kept within the home.

Contact and information sharing with healthcare practitioners was often undertaken by managers and office staff. This appeared a pragmatic approach given the time required, potential delays in reaching practitioners and the mobile nature of HCWs’ work, however, this could also isolate HCWs from communication across the wider care team:

> *When you’re a carer you don’t communicate with other community teams, it’s always the admins or care coordinators within the office….quite often the carer…is left quite isolated from that communication (3-HCW-8)*.

#### Inter-professional working can marginalise and isolate HCWs

Practitioners varied in their perceptions of HCWs. Some made positive statements about the value of their role and the importance of the HCW contribution:

> *I think they’re…often forgotten…you recognise that they’re there to do that certain role but actually they’re…the eyes and ears, aren’t they, really? (2-Practitioner-7)*.

However, others made disparaging comments, *it’s as if they’re not interested, that’s how I feel (1-Practitioner-1)*, and some HCWs and managers felt disregarded by practitioners who they believed did not value their views or expertise. Their roles did not always appear well understood or valued, and the everyday simplicity and familiarity of the care tasks they carried out (personal care, meal preparation) may have belied the depth of skills employed when supporting people at end-of-life.

They frequently referred to themselves as ‘just a homecare worker’. This was often a statement of how they believed others perceived them, with which they did not necessarily concur, *it would be nice if the carers are more recognised; we…do a great job (1-HCW-1).* However, others may have internalised this perspective and not recognised the significance of their contribution to client care.

Thus, it appears that HCWs can be marginalised and isolated within the wider care team, from whom they were often separated by hierarchical structures, poor communication and limited understanding of their roles. Analysis of HCW Pictor charts further emphasised HCW isolation, with practitioners, if present at all, often placed at a distance, whereas closer relationships with clients and carers were depicted.Where double-handed care was commissioned at end-of-life, the presence of a second HCW was often appreciated and could help mitigate a sense of isolation.

### Between case analysis

As noted in the methods section, the data from the three geographic sites was examined using matrix analysis to identify any significant differences. Overall, we found no substantial differences between the three areas, with differences more typically between individual agencies.The location of one homecare agency within a provider of statutory healthcare at Site Three facilitated inter-professional working, communication and information sharing, and represented a key difference for HCWs within that agency.There was considerable heterogeneity among agencies in respect of whether they had a specific focus on end-of-life care; the extent to which staff were offered end-of-life training; the support offered to HCWs after clients died; the minimum duration of calls provided; payment, including whether staff were reimbursed for travel time and costs; and the number of end-of-life clients at any time. However, this variation appeared between agencies rather than between sites. For example, within Site 3 homecare manager accounts of the care they provided differed markedly, and appeared to reflect the time available to HCWs, and the duration of the care commissioned (and accepted):

> *So it, it would just be your standard personal care, meal…preparation if they’re eating and drinking, medication support, toileting needs, but end-of-life in homecare is just very basic…it’s not that time to sit with family or with the person for the half an hour that…they may need you to sit and hold their hand (3-Manager-2)*
>
> *We would always fight for; I have to say….we’re not going in for half an hour calls…to sorta rush in, help somebody with some personal cares and then go out, because that’s all they’re commissioned for…. we always make sure that we have the right numbers of staff to support the service user and their family (3-Manager – 3)*.

There also appeared to be variation in care quality between agencies and among individual HCWs, suggesting that the provision of homecare at end-of-life may be patchy, and that the examples of positive and sensitive practice reported are not universal. Overall, homecare at end-of-life is subject to considerable variation, impacting on the experiences both of those delivering and in receipt of such care.

## Discussion

The study findings confirm the importance of homecare provision to clients at end-of-life and make an important contribution to understanding the experiences of HCWs which have received limited research and policy attention to date. Examining the findings through the lens of the adapted Ecological Systems Theory (Pask et al., 2018) helps capture the multiple relationships, systems and complexities which contribute to HCWs’ experiences of providing end-of-life care, with HCWs placed at the centre of the ecosystem for this analysis (Figure 1).

**Figure 1.**
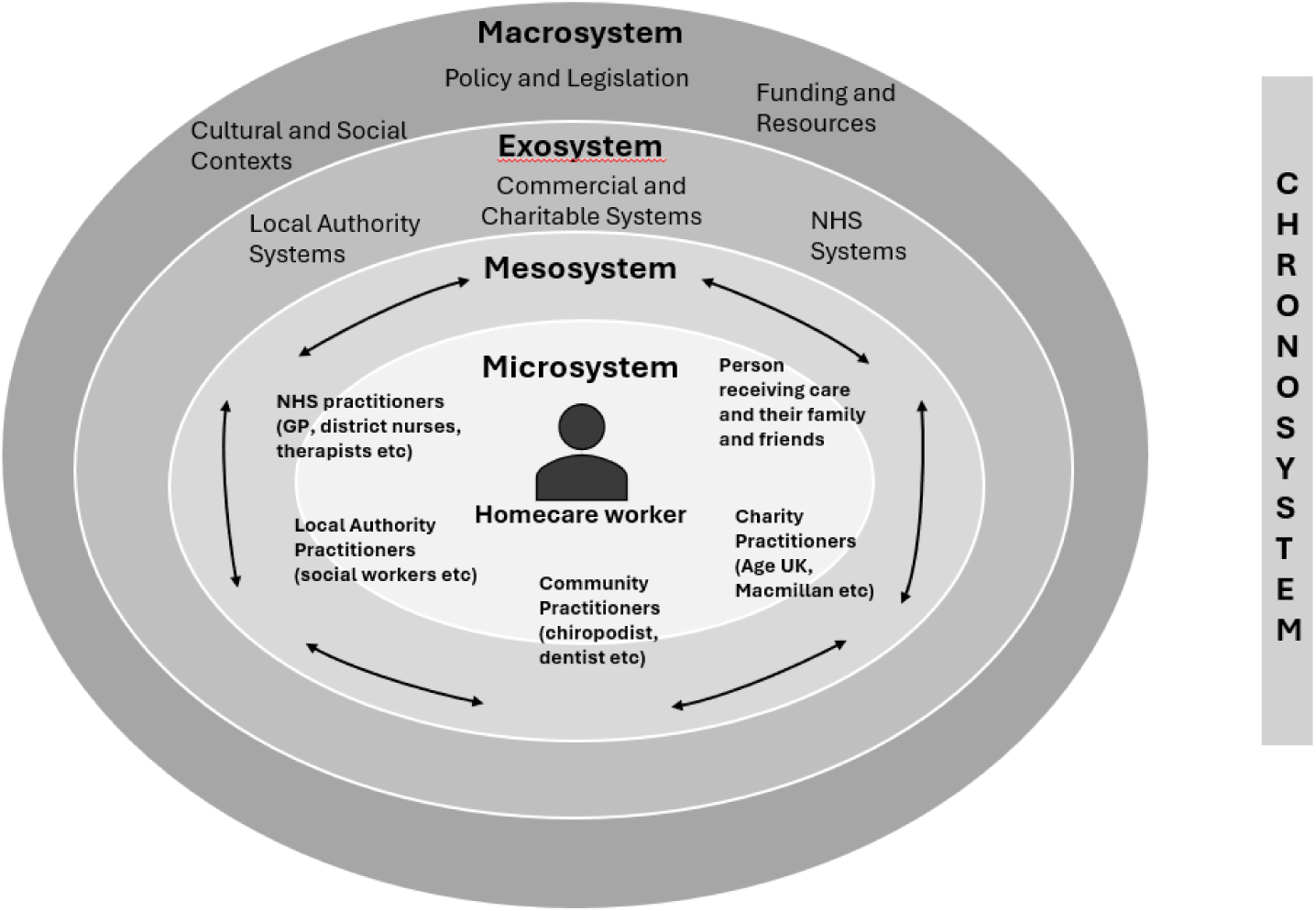
Eco-system homecare at end-of-life (adapted from Pask et al, 2018; Authors’ Own, under review).

### Microsystem – homecare worker needs and experiences

The practical and emotional workload for HCWs providing care at end-of-life forms part of the microsystem.

HCWs provide end-of-life care in a complex emotional and relational context.They are required to ‘read’ the emotional landscape and gauge the kind of care and interactions that are required as client and carer needs and preferences change, sometimes momentarily, requiring them to make nuanced and in-the-moment judgments. HCWs recognised the importance of attending to the details of individuals’ lives and the things that mattered to and comforted them, and supported clients to ‘live while dying’. The emotional climate in which HCWs deliver care is both challenging and impactful. Providing end-of-life care meant that death appeared an almost constant element of their lives while at work and beyond the workplace. HCWs experienced uncertainties which were underpinned by recognition of the unpredictability of the end-of-life trajectory, in which it is sometimes difficult to determine when individuals are approaching death (Mandelli et al., 2021). In this context their arrivals and departures from clients’ homes could be overshadowed by awareness that they might be first on the scene and/or making their final visit, and they appeared to anticipate death and their emotional responses to this. Consistent with other studies (Devlin and McIlfatrick, 2010; Abrams et al., 2019; Tsui et al, 2019; Findlay and Robertson, 2024), the emotional toll of supporting clients at end-of-life was documented, within which both unexpected and anticipated deaths could be experienced as distressing, and the loss of relationships with clients and carers was deeply felt, especially those of long duration.

### The mesosystem – relationships with clients, carers and practitioners

The mesosystem highlights the importance of relationships and interactions between HCWs, clients, carers and other practitioners. HCWs could develop close relationships with clients and carers (although examples of strained relationships were also given).These relationships were forged in the intimacy of care at end-of-life and were often experienced as akin to those of personal friends and families. HCWs often valued these relationships which were perceived as a cornerstone of good care, allowing HCWs to develop knowledge of individuals, their personalities and preferences, which informed the delivery of individualised, relational care. However, the closeness of these relationships could contribute to HCW distress, grief and bereavement.

The centrality of relationships with clients and carers also highlights the relevance of boundaries with clients, and with carers before and after death.There was potential for blurred boundaries in respect of requests to undertake tasks beyond the remit of their roles and training, and managing contact with bereaved carers after their relative/friend died, with HCWs required to develop relationships which were supportive and friendly, yet professional (see also Abrams et al, 2019). However, the findings draw attention to the difficulties of determining what appropriate boundaries in end-of-life relationships look like, and the challenges and ambiguities for HCWs in seeking to provide the depth of human care required, while not overstepping boundaries and appropriate professional behaviours. In considering ‘ambivalences’ within homecare for older people, Timonen and Lolich (2019, p737) highlighted the paradoxical expectation that:

### On the one hand, carers should be like family and friends, on the other hand, they must not get too close

HCWs need support and supervision to navigate the complexities of these potentially tangled emotional boundaries within the landscape of good care provision. However, manager responses indicated that they could themselves experience difficulties defining appropriate boundaries and, in some instances, turned a ‘blind eye’ when they suspected that some went over and beyond accepted behaviours, such as maintaining protracted contact with bereaved carers.

The mesosystem also incorporates the sometimes more distal, but important relationships and interactions with members of the wider health and social care network around the person approaching end-of-life.The findings indicated that HCWs and practitioners may develop positive relationships in which their mutual roles are valued. However, it was evident that HCWs could be marginalised within the care team; their role poorly understood and valued, and the skills and inherent complexity within their role unrecognised, as highlighted elsewhere (Manthorpe et al., 2019; Forward et al 2024). Further, they may be left out of communication and information sharing, and feel disregarded.This contrasts with the view that communication is central to good practice, and in its absence care may be compromised if HCWs are not able to share their insights which are informed by close and regular contact with clients (Forward et al., 2024).

### Exosystem – wider care arrangements

The need for care which considers the needs of carers as well as clients was evident to HCWs, but not always well reflected in commissioning practices which defined their roles with individual clients. Therefore, support for carers who provide considerable end-of-life care, but who often also need practical assistance and emotional support (Ewing et al, 2012; Mogan et al, 2024) may be squeezed into already tightly timed calls, with the risk of amplifying carer isolation and distress.

The ways in which care was commissioned and services designed could contribute to HCWs’ experiences at end-of-life. Although our analysis indicates that much variation in care delivery is at the level of individual workers and agencies, there were local design/commissioning decisions which influenced homecare practices and experiences. For example, locating a homecare provider within a provider of statutory healthcare appeared to enhance working relationships, communication and information sharing between HCWs and health practitioners. Further, commissioning practices in respect of out-of-hours palliative care services impact on the extent to which clients and carers can access crisis support (Firth et al, 2025), with consequent benefits (or challenges) for the HCWs supporting them.The study highlighted the varying models of end-of-life homecare provision, including agencies which advertised provision of end-of-life care alongside others which positioned themselves as providers of generic care only (yet which included caring for clients when dying).

Research to identify the benefits (and drawbacks) of different models of delivery, in respect of client care and HCW experiences, training and support, may make a valuable contribution to understanding the sector and optimal approaches to end-of-life care.

### The macrosystem – cultural, political and societal influences

The macrosystem emphasises the societal and political factors which influence care provision and the HCW experience. Social care has long been overlooked by policy makers, and public perceptions of social care are poor (Authors’ Own (a) – in preparation). Homecare has received scant attention within end-of-life care policy, meaning there is a limited policy impetus and attention to the development of support and training for HCWs at end-of-life (Authors’ Own (a) – in preparation).

Other policy directives may inadvertently contribute to challenges within the sector. For example, there is increased use of digital records within homecare (Healey et al., 2024) reflecting a strong policy drive towards digitisation within social care (Department of Health and Social Care, 2021). However, the incompatibility of systems within health and social care may contribute to fragmented communication and information sharing across organisational boundaries, which may hinder inter-professional working, in comparison to the use of easily accessible paper-based systems.The UK Government’s announcement of the development of a shared digital platform for NHS and care staff (Department of Health and Social Care, 2025) may help alleviate this issue.The Government has also signalled an expectation that social care workers may be required to assume further responsibilities and undertake tasks (such as checking blood pressure) to relieve pressures on the health service (Department of Health and Social Care, 2025).These may further stretch homecare workers and the social care sector as a whole, despite being poorly resourced in comparison to the health service, and further suggesting a devaluing of the distinct role of social and non-medical care, and an underpinning assumption that those who are ‘just a homecare worker’ can readily absorb additional roles.

### Chronosystem – the dynamic influence of time

Supporting clients at end-of-life required HCWs to be flexible and responsive to often rapidly changing needs and fluctuations, in which practical tasks mandated on care plans were not always required during the visit or were undertaken during a later call when clients were more alert and able to tolerate care.This reflects the findings of Abrams et al., (2019) in which HCWs identified adaptability and flexibility as one of their key contributions during end-of-life care. Clients and carers required and valued emotional support and information provision, and these were often perceived as the priority, although they were seldom on the care plan. Thus, while care plans could be informative, they did not always represent clients’ changing needs and had to be adapted in practice. Further, in addition to familiar practical care tasks, new and less familiar tasks are required, both as clients approach end-of-life and at the time of death. In our increasingly diverse society, different cultural beliefs inform client and carer needs before and after death (Hossain et al, 2020); the ability to respond to these represents a further element of flexibility required of HCWs.

The imbalance between the time allocated for care calls and the time required to meet clients’ and carers’ needs has been an ongoing issue in homecare at end-of-life and more generally (Devlin and McIlfatrick, 2010; Burns et al., 2023; Strandell, 2023).Where brief visits are commissioned, emotional and carer support must be accommodated alongside practical tasks, care may be rushed, and HCWs’ abilities to stay on if additional support is needed or to respond to crises, conflicts with the need to meet obligations to other clients (Strandell, 2023).Writing about homecare for older people Strandell (2023, 213) observes that *unpredictable situations happen more or less daily*, however, it might be expected that in end-of-life care this unpredictability is amplified, deepening time-pressures and the need for flexibility. In this climate, responding to needs not on the care plan often relied on HCW goodwill (Devlin and McIlfatrick, 2010; Strandell, 2023), with workers ‘going beyond’ to meet clients’ needs from their own resources (temporal, personal or financial), despite their own low pay and time-pressured working conditions.Thus, the political influences at the macrosystem and commissioning practice at the exosystem, impact on the chronosystem creating an imbalance between the level of client and carer needs and the availability of funded time. In these circumstances the responsibility *to reconcile the tensions inherent in a care plan that offers no time to care* (Bolton and Wibberley, 2014, p689) is shouldered by the least well-resourced members of the care network, while providing little incentive for wider systemic change (Strandall, 2023). However, where service design and commissioning practices meant that brief or timed calls were not undertaken, HCWs valued working conditions which furnished them with greater flexibility to respond to changing client and carer needs, and respond to people’s emotional and social needs without rushing.

## Strengths and limitations

This study brought together the perspectives of multiple stakeholders in the currently under-explored area of homecare delivery at end-of-life, broadening attention from a recent UK research focus on homecare at end-of-life in the context of dementia care (Yeh et al., 2018; Abrams et al., 2019; Manthorpe et al, 2019). Participants worked with or received care from a wide range of homecare agencies in three geographic areas, meaning that there was considerable variation in policies and practices between agencies and localities, and that a wide range of experiences of homecare were captured. Data collection and analysis was undertaken by a team of researchers with varied professional backgrounds and including team members bringing public perspectives, allowing for a robust analysis incorporating multiple perspectives.

We acknowledge some study limitations. Although some recruitment was independent of agencies, our HCW recruitment was frequently facilitated by managers who, as reported elsewhere (Manthorpe et al, 2019), may have screened out staff they believed would provide negative or critical commentaries, and the agency may have declined to participate if concerned about the quality of care provided or likely staff responses. Although some HCW participants were recruited directly, the perspectives of those delivering care in particularly challenging agency and organisational contexts may have been under-represented.Therefore, the HCW and manager accounts often provide evidence of best practice; this contrasts with the experiences of some participants and the wider literature in which accounts of poor and inadequate care practices at the individual or agency level have been reported (Martinsen et al, 2024;Tanner et al, 2024) highlighting the diversity in quality of end-of-life care. Our data however illustrates the value of homecare worker support and the depth and sensitivity of the practical and emotional support which can be delivered when HCWs are well supported, trained and informed about their role and responsibilities, and when external factors impact on social care support the recruitment and retention of HCWs. Our research has also explored the training and support needs of HCWs supporting clients at EOL (Authors’ Own (b) – in preparation), attention to these may enable HCWs to be better equipped to support clients, and foster their wellbeing in a context of uncertainty and emotional demands.

The majority of HCW participants were White British, although care workers from other cultural groups were included; all clients and all but one carer were White British. James et al (2024) have highlighted the importance for homecare recipients of having HCWs with whom they have a shared language, and who know how to prepare their preferred food and drink; our study also identified challenges in the absence of shared language. Further research is needed to explore these challenges, how they are experienced and how HCWs, clients and carers manage gaps in respect of language and understanding of how to provide care which is tailored to individual and cultural preferences.

Lastly, only about a third opted to use a Pictor guided interview, at least in part due to the preference of many to have on-line interviews. However, as the study design was not dependent on this method, we were able to generate a rich dataset which was strengthened by the Pictor charts that were made, particularly illustrating clearly the isolation HCWs experienced. It would be interesting in future research to conduct a Pictor-only study employing face-to-face interviews.

## Conclusion

This paper presents findings from a study examining the experiences of HCWs providing homecare for those approaching end-of-life.The interviews involved multiple stakeholders and gave rich insights into the challenges and privileges experienced by HCWs in this context.The themes presented explore the range of experiences and shed light on potential support needs for this group. By discussing the findings in the context of the adapted Bronfenbrenner’s Ecological Systems Theory, aspects of the data can be considered at the different levels of the systems of care which support people approaching end-of-life in the community. A systemic approach to data analysis has enabled and deepened an understanding of the complexity of the roles of HCWs, the pressures they experience and the social contexts within which they work. They sit at the heart of the delivery of end-of-life care in the home yet are excluded from systems which surround them – of communication, decision making, social acknowledgement and financial reward. The implications of this study need to be considered at practice, research and policy levels, if the value and impact of HCWs in providing end-of-life care is to be realised.

## Data availability

Access to our data can be made available on request by authorised researchers following completion of a data sharing agreement.To request access, contact the study authors, or citing Worktribe Output ID 5179695.

## Acknowledgements

We extend our thanks to all those who supported this study.This includes homecare organisations, Health Trusts, and hospices, who facilitated participant recruitment; homecare workers, managers, clients, carers, health and social care practitioners and commissioners who participated in research interviews; our homecare worker and service user and carer advisory groups.Their support was invaluable, and we thank them all for their contribution to the research.

We also extend our thanks to Kathryn Harvey our Research Study Administrator who has ably supported with many practical aspects of the study and enabled us to bring this study to fruition.

## Appendices

**Table 1.** Inclusion/exclusion criteria for each participant group.

| Participant Group | Inclusion Criteria | Exclusion Criteria |
| --- | --- | --- |
| Homecare workers | Provide EOL care as part of their role, or have provided such care within the previous 12 months for one or more clients. | Experience of EOL care was not within the previous 12 months. |
| Homecare managers | Managers who provide homecare (including EOL homecare) , or who have had this role within the previous 12 months. | Have not provided homecare within the past 12 months at EOL. |
| Homecare clients | Individuals receiving homecare at EOL and who are considered by the care provider to be in the last 6 months of life.<br>Have capacity to consent to participation (verbal or written).<br>Are able to participate in an interview lasting 30 – 60 minutes.<br>Have sufficient English communication skills to participate in an interview, or are able to participate with support from a communication assistant/interpreter. | Are anticipated to be likely to die within the next 7 days (by homecare providers or a health practitioner). |
| Carers | Are a family member or friend of a client at EOL who is receiving homecare (currently or previously).<br>If former carers to be at least 3 months, and no more than 12 months, post-bereavement.<br>Have sufficient English communication skills to participate in an interview, or are able to participate with support from a communication assistant/interpreter. | A carer of a person anticipated to die within 7 days as reported by a homecare provider or health practitioner.<br>If a former carer, at least 3 months, and no more than 12 months, post-bereavement. |
| Health and social care practitioners | Health and social care practitioners who support clients at EOL or who have done so in the past 12 months. This includes GPs, social workers, community and district nurses. | Practitioners who have not supported people at EOL (i.e. within the previous 12 months). |
| Commissioners | Working to commission homecare, including EOL care, or who have had this role in the previous 12 months. | Have not had a role in commissioning homecare, including homecare at EOL in the previous 12 months. |

**Table 2.**
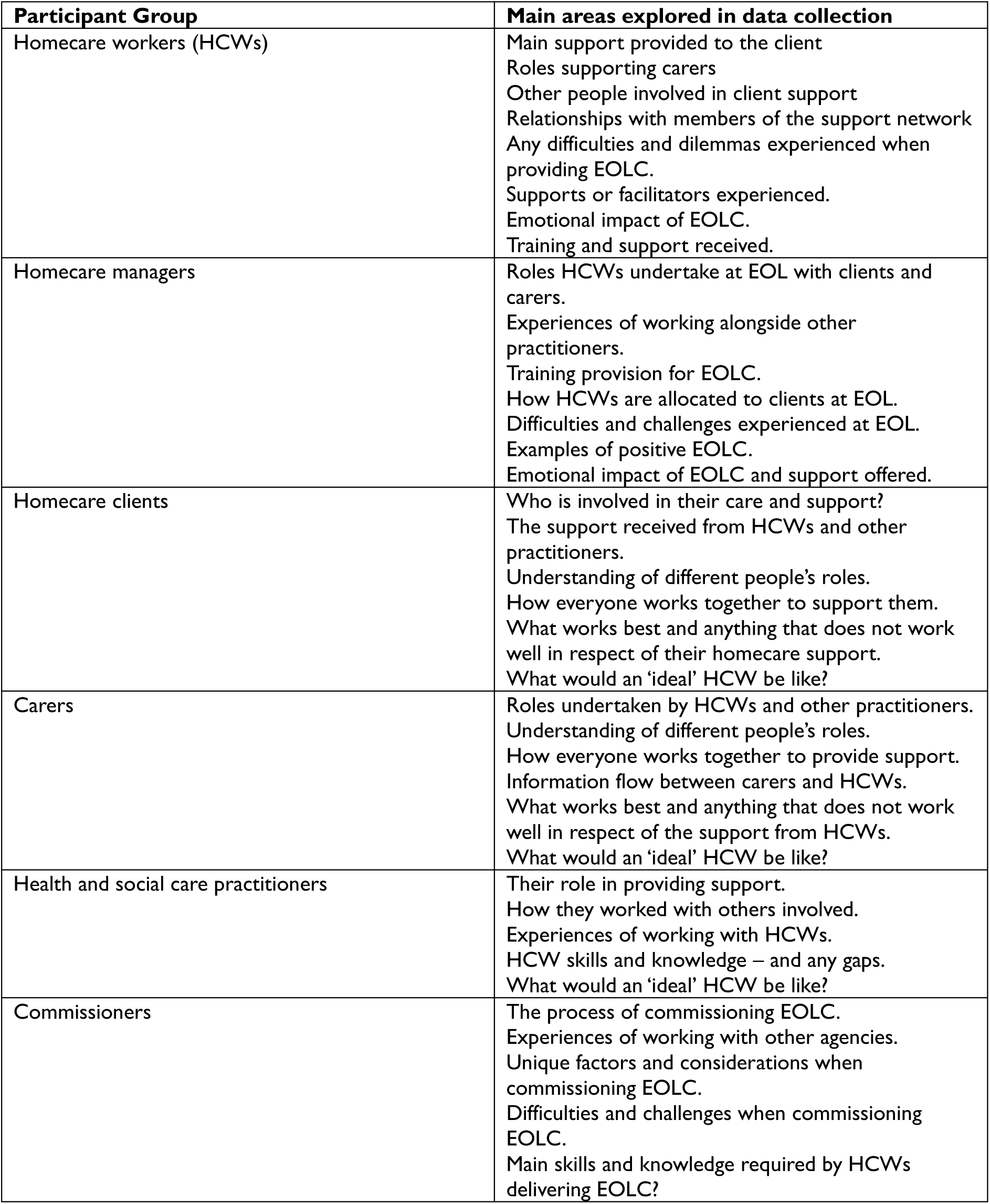
Key questions explored in interviews for each participant group.

| **Participant Group** | **Main areas explored in data collection** |
| --- | --- |
| Homecare workers (HCWs) | Main support provided to the client<br>Roles supporting carers<br>Other people involved in client support<br>Relationships with members of the support network<br>Any difficulties and dilemmas experienced when providing EOLC.<br>Supports or facilitators experienced.<br>Emotional impact of EOLC.<br>Training and support received. |
| Homecare managers | Roles HCWs undertake at EOL with clients and carers.<br>Experiences of working alongside other practitioners.<br>Training provision for EOLC.<br>How HCWs are allocated to clients at EOL.<br>Difficulties and challenges experienced at EOL.<br>Examples of positive EOLC.<br>Emotional impact of EOLC and support offered. |
| Homecare clients | Who is involved in their care and support?<br>The support received from HCWs and other practitioners.<br>Understanding of different people's roles.<br>How everyone works together to support them.<br>What works best and anything that does not work well in respect of their homecare support.<br>What would an 'ideal' HCW be like? |
| Carers | Roles undertaken by HCWs and other practitioners.<br>Understanding of different people's roles.<br>How everyone works together to provide support.<br>Information flow between carers and HCWs.<br>What works best and anything that does not work well in respect of the support from HCWs.<br>What would an 'ideal' HCW be like? |
| Health and social care practitioners | Their role in providing support.<br>How they worked with others involved.<br>Experiences of working with HCWs.<br>HCW skills and knowledge – and any gaps.<br>What would an 'ideal' HCW be like? |
| Commissioners | The process of commissioning EOLC.<br>Experiences of working with other agencies.<br>Unique factors and considerations when commissioning EOLC.<br>Difficulties and challenges when commissioning EOLC.<br>Main skills and knowledge required by HCWs delivering EOLC? |

